# Association between environmental pollution and prevalence of coronavirus disease 2019 (COVID-19) in Italy

**DOI:** 10.1101/2020.04.22.20075986

**Authors:** Giuseppe Lippi, Fabian Sanchis-Gomar, Brandon M. Henry

## Abstract

The novel coronavirus disease 2019 (COVID-19) has recently been upgraded to a pandemic by the World Health Organization due to the alarming levels of spread and severity. Since several lines of evidence also attest that Lombardy region has an extraordinarily high level of environmental pollution, we aimed to explore the potential epidemiological association between the number of cases of COVID-19 and environmental pollution in Italy. Data on environmental pollution in Italy were retrieved from the 2019 annual report of the organization Legambiente (League for the Ambient). The adjusted correlation between the number of days in which environmental pollutants exceeded established limits and the overall number of COVID-19 cases reveals the existence of a highly significant positive association (r=0.66; 95% CI, 0.48-0.79; p<0.001). The association remained statistically significant even when the number of days above pollutant limits was correlated with the number of COVID-19 cases per 1000 inhabitants (r=0.43; 95% CI, 0.18-0.62; p=0.001). Living in a province with over 100 days per year in which environmental pollutants were exceeded was found to be associated with a nearly 3-fold higher risk of being positive for COVID-19 (0.014 vs. 0.005 COVID-19 cases per 1000 inhabitants; OR, 2.96; 95% 2.12-4.13; p<0.001). Reinforced restrictive measures shall be considered in areas with higher air pollution, where the virus is more likely to find a fertile biological or environmental setting.

## 1. Introduction

The novel coronavirus disease 2019 (COVID-19) has recently been upgraded to a pandemic by the World Health Organization (WHO), due to the alarming levels of spread and severity (Mahase, 2020). Though nearly all countries across the globe have now been virtually involved, the burden of disease seems to progress heterogeneously among different countries and territories. For example, Remuzzi and colleagues recently highlighted that some parts of Italy, especially the Lombardy region, are experiencing a considerably vaster outbreak compared to other areas of the country (Remuzzi and Remuzzi, 2020). Since several lines of evidence also attest that this Italian territory has an extraordinarily high level of environmental pollution (Carugno et al., 2017), we aimed to explore the potential epidemiological association between the number of cases of COVID-19 and environmental pollution in Italy.

## 2. Methods

Data on environmental pollution in Italy were retrieved from the 2019 annual report of the organization Legambiente (League for the Ambient), an association which annually pools information published by all Regional Agencies for Environmental Prevention and Protection in Italy (https://www.legambiente.it/wpcontent/uploads/Malaria2019_dossier.pdf). The extent of pollution within each specific Italian province was expressed in terms of days per year during which the limits set for particular matter 10 (PM10; 50 μg/m^3^ daily) or ozone (120 μg/m^3^ in 8 hours) were exceeded (available data were limited to Italian provinces with ≥26 days of limits exceeded). Data on the overall number of positive COVID-19 cases per province was captured from the daily statics made available by the Italian Ministry of Health, on its official website, as of April 5, 2020 (http://www.salute.gov.it/imgs/C_17_pagineAree_5351_16_file.pdf). The association between local environmental pollution and number of positive COVID-19 cases officially reported by the Italian Ministry of Health was then analyzed by considering the number of days per year during which the limits of PM10 and ozone had been exceeded as both a continuous (i.e., using linear regression analysis and Pearson’s correlation) and categorical (i.e., by calculation of the odds ratio; OR) variable. Statistical analysis was carried out with Analyse-it (Analyse-it Software Ltd, Leeds, UK). The study was performed in accordance with the Declaration of Helsinki, under the terms of relevant local legislation.

## 3. Results

The adjusted correlation between the number of days in which environmental pollutants exceeded established limits and the overall number of COVID-19 cases as recorded by the Italian Ministry of Health is shown in Figure 1a, which reveals the existence of a highly significant positive association (r=0.66; 95% CI, 0.48-0.79; p<0.001). The association remained statistically significant even when the number of days above pollutant limits was correlated with the number of COVID-19 cases per 1000 inhabitants (r=0.43; 95% CI, 0.18-0.62; p=0.001) (Figure 1b). Importantly, living in a province with over 100 days per year in which environmental pollutants were exceeded was found to be associated with a nearly 3-fold higher risk of being positive for COVID-19 (0.014 vs. 0.005 COVID-19 cases per 1000 inhabitants; OR, 2.96; 95% 2.12-4.13; p<0.001).

**Figure 1.**
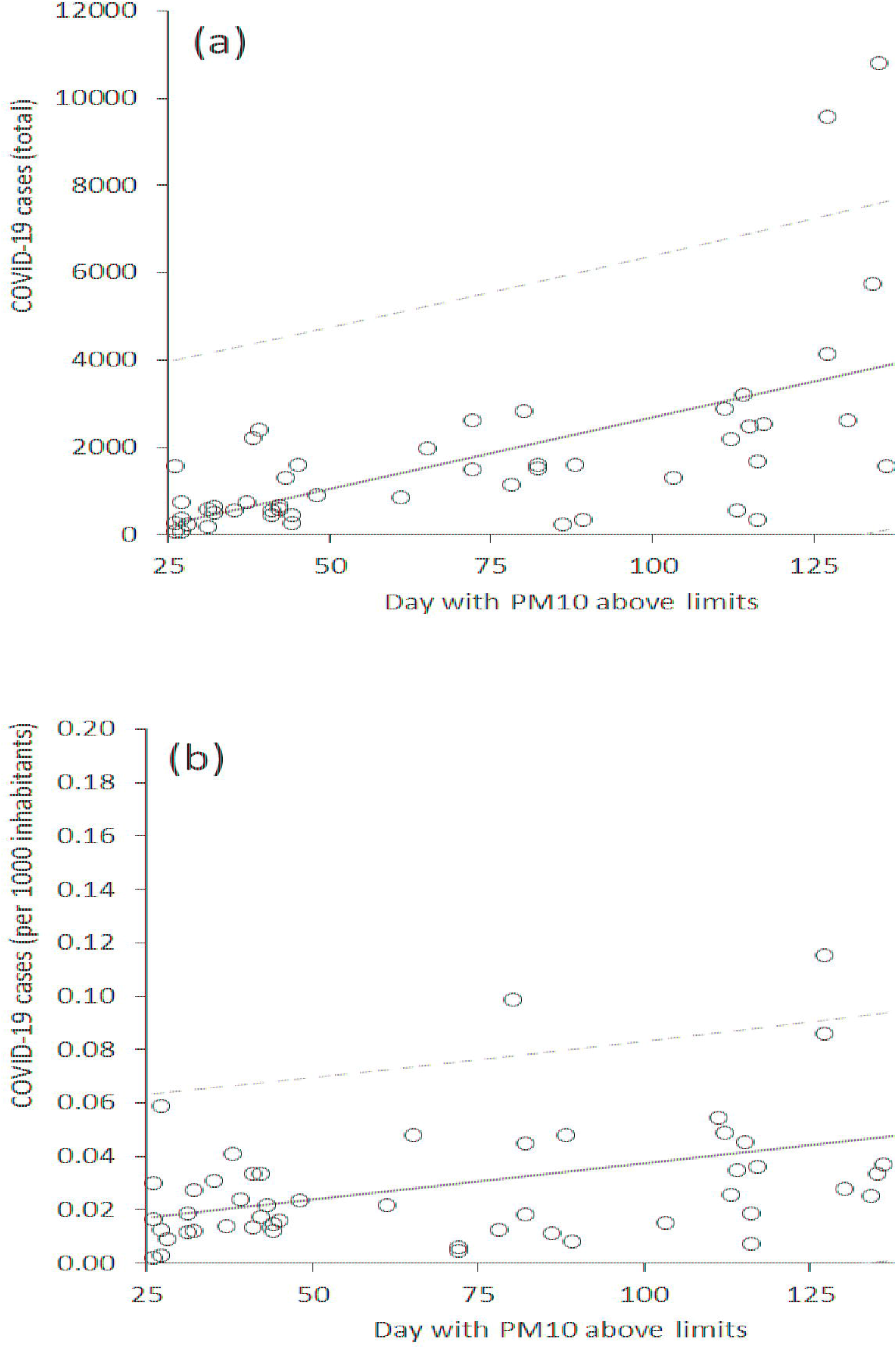
Correlation between environmental pollution and coronavirus disease 2019 (COVID-19), expressed as either total number of COVID-19 cases (a) or number of COVID-19 cases per 1000 inhabitants (b).

## 4. Discussion

The results of this early analysis support the existence of a significant correlation between the number of COVID-19 cases and environmental pollution in Italy. Although we cannot rule out that this association may be mostly casual, some reliable biological explanations would support a direct causal relationship. Severe acute respiratory disease coronavirus 2 (SARS-CoV-2), the microorganism responsible for COVID-19, is a respiratory virus, which mostly infects type 2 pneumocytes, causing a severe form of interstitial pneumonia, occasionally evolving to acute respiratory distress syndrome (ARDS) (Remuzzi and Remuzzi, 2020). Previous studies have highlighted the existence of a significant association between air pollutants exposure and enhanced risk of developing respiratory viral infections (Ciencewicki and Jaspers, 2007), such that there is no reason to think that COVID-19 will be an exception. Moreover, according to National Aeronautics and Space Administration (NASA), China’s air pollution has significantly dropped during the coronavirus outbreak, with carbon emissions dropping by 25% (https://earthobservatory.nasa.gov/blogs/earthmatters/2020/03/05/howthe-coronavirus-is-and-is-not-affecting-the-environment/). These decreases in air pollution parallel the decline in the number of cases in that country. Interestingly, the Copernicus Sentinel-5P satellite of the European Space Agency (ESA) has also detected a dramatic decline in air pollution over northern Italy following lockdown (https://www.esa.int/ESA_Multimedia/Videos/2020/03/Coronavirus_nitrogen_dioxide_emissions_drop_over_Italy). Hopefully, this will correspond to a definitive decline in the number of cases.

## 5. Conclusion

Our findings support the concept that reinforced restrictive measures shall be considered in areas with higher air pollution, where the virus is more likely to find a fertile biological or environmental setting.

## Data Availability

The data that support the findings of this study are available from the corresponding author,FSG, upon reasonable request.

## Acknowledgments

Fabian Sanchis-Gomar is supported by a postdoctoral contract granted by *“Subprograma Atracció de Talent - Contractes Postdoctorals de la Universitat de València”*.

